# Epidemiology of pediatric cancer in the Northwest Region of Cameroon

**DOI:** 10.1101/2020.02.04.20020487

**Authors:** Rohit Borah, Francine Kouya

## Abstract

**Background:** The epidemiological scale of cancer in Cameroon is still relatively unknown, as public health research in sub-Saharan Africa has traditionally focused on infectious and/or communicable disease. As awareness for chronic disease such as cancer has risen, so, too, have diagnoses and incidence. In order to ensure more effective care and quality of life for children at greater risk for cancer diagnoses, specific and effective research must be conducted in order to institute effective policy and protocols.

**Objective:** To comprehensively review all pediatric cancer cases seen in the Northwest Region of Cameroon in order to provide novel, current epidemiological data on pediatric cancer in the region.

**Methods:** The authors retrospectively reviewed each individual pediatric cancer case seen from January 1, 2016, through November 30, 2017, in the three largest hospitals in the Northwest Region of Cameroon: Mbingo Baptist Hospital, Bamenda Provincial Hospital, and Banso Baptist Hospital.

**Results:** 173 cases of pediatric cancer were identified as being treated in one of the three hospitals in question over the 25-month study period. The average age of the patients was 6.23 years (SD = ± 3.93), and 58.4% of the patients were male. The three most common types of cancer diagnosed in these pediatric patients were Burkitt lymphoma, retinoblastoma, and Wilms tumor. Of the 173 cases, 105 of the patients were originally from the Northwest Region. The incidence of pediatric cancer originating from the Northwest Region of Cameroon is 5.92 cases (95% CI: 4.79 – 7.05) per 100,000 persons per year.

**Conclusion:** This study is the first extensive examination of pediatric cancer epidemiology in the Northwest Region of Cameroon. Children in sub-Saharan Africa are disproportionately affected by cancer in comparison with their counterparts in the developed world. As the body of literature continues to grow in the years to come, more effective care both preventive and curative can affect the lives of millions.

## Introduction

Cancer and other chronic disease profoundly affect the developing world with high incidence, prevalence, and mortality rates. Public health research and practitioners have focused for decades on the challenges presented by infectious and/or communicable diseases such as malaria, tuberculosis, and HIV/AIDS. Due both to greater awareness of cancer and improving resources devoted to care, the true burden of cancer, both pediatric and adult, is now more effectively being treated and studied across Africa.

The National Cancer Institute of the United States defines pediatric cancers as “cancers that occur between birth and 15 years of age.^1^” Childhood cancers are more prevalent in sub-Saharan Africa than in high-income countries: in sub-Saharan Africa, pediatric cancers account for 4.6% of all cancers in comparison to 0.5% in high-income countries.^2^ As awareness and education improve, an increase in the number of reported cases is expected. This increase can improve the quality of treatment, the comprehensiveness of registries, and the overall effectiveness of the cancer care environment across the continent.

In Cameroon, public health attention has shifted towards cancer treatment and prevention over the past decade. However, the true epidemiological burden of cancer in Cameroon is relatively unknown as there is little data available.^3^ A 2007 study commissioned by the Cameroonian National Cancer Committee (CNCC) determined that lymphomas, specifically Burkitt lymphomas (BL), are the most prevalent childhood cancer in the country.^4^ The most comprehensive study of cancer epidemiology in Cameroon was conducted at General Hospital Yaoundé, the capital city, in 2012. This study examined cancer incidence across all age groups in Yaoundé and found the age-standardized cancer rate for all cancers to be 53.35 cases per 100,000 people.^5^ However, that research is not directly translational to this study due to the ethnic, environmental, and geographical differences between the Yaoundé and the Northwest Region.

The previous study most applicable to this research examined the epidemiology of Burkitt lymphoma in the Northwest Region from 2003 to 2010.^6^ The objective of that research was to determine if a relationship exists between malarial infection and BL. While no relationship was demonstrated, the researchers found that the average yearly incidence rate of BL from 2003 to 2010 in the Northwest Region was 2.57 per 100,000 children.

Developing countries across the world have seen incident rates of pediatric cancer rise over the past decade due to improved education and training for medical personnel leading to more diagnoses. Updated, comprehensive epidemiological research is necessary in order to more effectively understand the pervasiveness of the pediatric cancer problem.

The primary purpose of this study was to update and complete the epidemiological data on pediatric cancer in Northwest Cameroon. The study aimed to determine the overall incidence of pediatric cancer in the Northwest Region of Cameroon in 2016-2017 and compare that incidence to previous studies of cancer epidemiology in Cameroon.

## Methods

This study examined new cancer cases presented in the Northwest Region of Cameroon from January 1, 2016, through November 30, 2017. The patient data was collected from the three largest hospitals in the region: Mbingo Baptist Hospital (MBH), Bamenda Provincial Hospital (BPH), and Banso Baptist Hospital (BBH). These are the only three hospitals equipped to treat pediatric cancer. Other, smaller hospitals in the region such as Shisong Hospital typically refer potential cancer patients to one of these three hospitals.

### Data synthesis

Demographic data was collected by Rohit Borah (RB), a public health researcher trained at the University of Alabama at Birmingham in Birmingham, AL, U.S.A. The following demographic data was collected: patient name, age, gender, height, weight, the hospital at which the patient was treated, and admission date.

Treatment and outcome data was collected jointly by Dr. Francine Kouya, the pediatric oncology specialist at MBH, and RB. This data comprises the type of cancer diagnosed, the stage at which the patient was diagnosed, the length of time for which the patient experienced symptoms, the treatment administered, and the inpatient outcome.

At MBH, the patient information is primarily kept in physical patient files with handwritten physicians’ and nurses’ notes. The oncology data, specifically cancer type and staging, was taken from pathologists’ reports and physicians’ notes. Both authors reviewed each patient file, and any discrepancies were solved via arbitration. The patient information from BPH was collected in a ledger, and specific oncology information for each patient was given by nurses.

### Data management and analysis

All statistical coding, organization, and analyses were conducted using Microsoft Excel and R, a programming language for statistical computing and graphics.^7^ Descriptive statistics for the variables in question and the key performance indicators were tabulated via R. The graphs, charts, or graphics created solely for the purpose of this research were created via either Tableau, a data visualization software, or the “ggplot2” R package.^8^

The primary data this research aimed to provide is the incidence for pediatric cancer in Northwest Cameroon. The United States Centers for Disease Control and Prevention defines incidence as “the occurrence of new cases of disease or injury in a population over a specified period of time.^9^” Incidence was determined via the following equation: 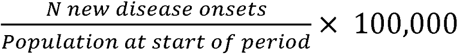. In order to accurately assess the pediatric population for the Northwest Region, the researchers extrapolated data from the 2010 Cameroonian national census and the 2016 population estimate of the United States Central Intelligence Agency’s World Factbook.^10,11^ The expected pediatric population for the Northwest Region of Cameroon in 2016 was 932,370. These calculations can be found in **Table 1**.

**Table 1:** Cameroon Population Statistics

|  | Total Population | % Of Total | Pediatric Population | % Of Total |
| --- | --- | --- | --- | --- |
| **Cameroon (2010)** | 19,406,100 | - | 8,465,364 | 43.62% |
| **Cameroon (2016)** | 23,130,708 | - | 9,929,106 | 42.93% |
| **Northwest (2010)** | 1,804,695 | 9.30% | 776,113* | 43.01% |
| **Northwest (2016)** | 2,151,156* | 9.30% | 925,212* | 43.01% |

\*Estimated population parameter based on YoY population growth

Missing values were present for several variables. The primary variables of interest were hospital at which treatment was administered, admission year, patient sex, cancer diagnosis, and patient region of origin. Any data with missing values for cancer diagnosis, admission year, or hospital of treatment were subject to listwise deletion. Missing values for both patient region of origin and patient sex were determined to be missing completely at random, due to improper reporting. The authors used conditional mean imputation to substitute the missing values and conduct the analysis.^12^ For the descriptive statistics regarding the other variables such as staging and patient height and weight, no imputation was used.

### Patient and Public Involvement

This study was conducted as a retrospective observational examination of health data over the preceding 23 months. Protocol approval was granted on December 19, 2018, by the Cameroon Baptist Convention Institutional Review Board to collect relevant patient information. This permitted collection of demographic, treatment, and outcome data for pediatric cancer patients seen for the period of January 1, 2016, to November 30, 2017. Each patient was assigned a unique numeric identifier for the purpose of this study in order to ensure that the patients’ identities and health information remained secure and private as per the IRB approval.

## Results

173 cases of pediatric cancer were identified from the period January 1, 2016, to November 30, 2017. The median age was 5.75 years with a mean of 6.23 years (SD ± 3.93). 101 (58.4%) of the patients were male, and 72 (41.6%) were female. Three of the 173 patients have only reported demographic data. Those patients are excluded from the following analyses apart from incidence.

There were 26 different types of cancers diagnosed over the course of study. The three most common types of cancer were Burkitt lymphoma (31.2% of all diagnoses), retinoblastoma (19.1%), and Wilms tumor (16.2%). **Figure 1** shows the treatment outcome by diagnosis for the seven most commonly diagnosed cancers in the Northwest Region of Cameroon. **Figure 2** breaks down the ages of diagnosis for these most common cancers. Patients with unreported outcomes are not shown in these graphics. The full diagnosis data is listed in **Table 2**. These findings are consistent with 2015 research of cancer registries in countries across sub-Saharan Africa.^13^ That study found that “unlike developed countries, lymphomas, nephroblastoma [also known as Wilms’ tumor], Kaposi sarcoma, and retinoblastoma were the most common pediatric tumors in Africa.”

**Figure 1:**
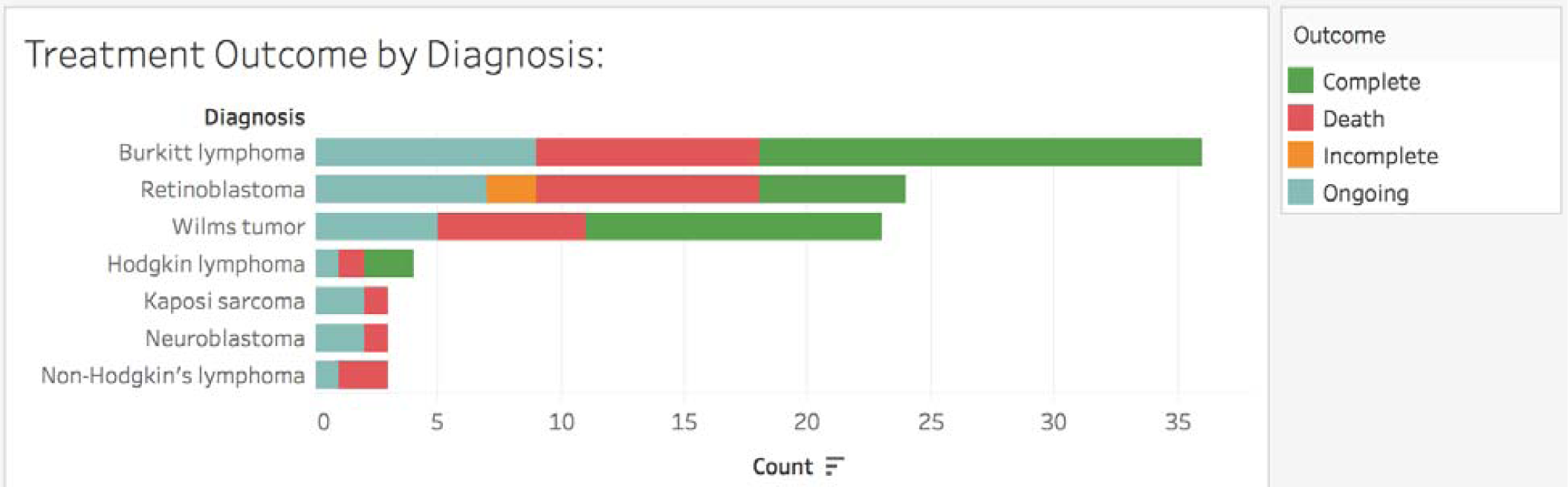
Treatment outcome for the most commonly diagnosed cancers.

**Figure 2:**
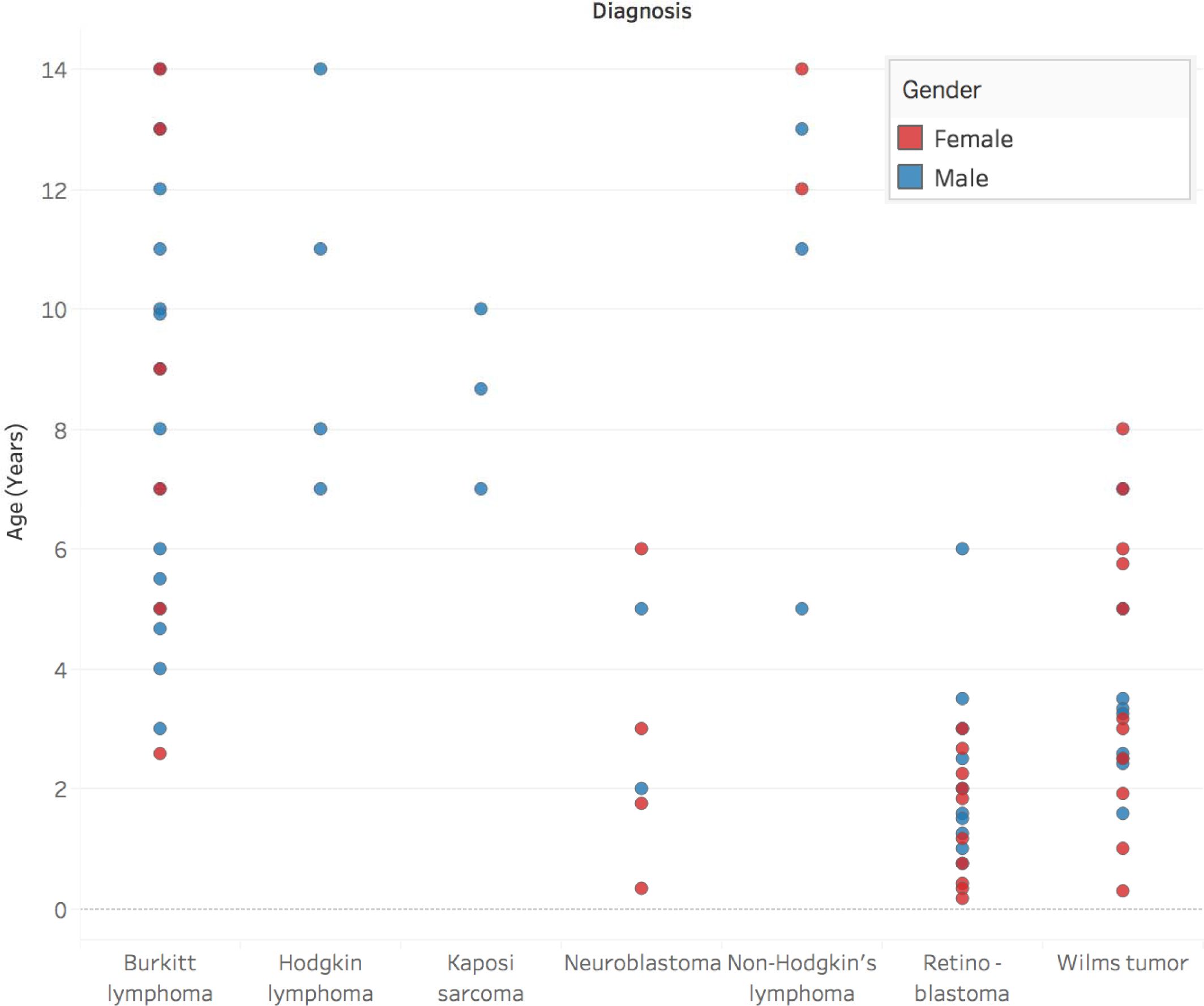
Age by diagnosis for the most commonly diagnosed cancers.

**Table 2:** Types of Cancer Diagnosed by Gender

| Diagnosis | Female: | Male: | Total: |
| --- | --- | --- | --- |
| Acute myeloid leukemia |  | 2 | 2 |
| ALL | 1 | 1 | 2 |
| Atrial Myxoma |  | 1 | 1 |
| Brain tumor | 1 | 1 | 2 |
| Burkitt lymphoma | 17 | 37 | 54 |
| Cerebral lymphoma |  | 1 | 1 |
| Colon carcinoma | 1 |  | 1 |
| Hepatoblastoma | 1 |  | 1 |
| Hodgkin lymphoma |  | 4 | 4 |
| Kaposi sarcoma | 1 | 3 | 4 |
| Langerhans cell histiocytosis |  | 1 | 1 |
| Leukemia | 1 | 1 | 2 |
| Lymphoblastic leukemia | 2 |  | 2 |
| Lymphoma | 1 |  | 1 |
| Nasopharyngeal Carcinoma |  | 2 | 2 |
| Neuroblastoma | 4 | 3 | 7 |
| Non-Hodgkin's lymphoma | 4 | 6 | 10 |
| Osteochondroma | 1 |  | 1 |
| Osteochroma | 1 |  | 1 |
| Osteosarcoma |  | 3 | 3 |
| Retinoblastoma | 17 | 16 | 33 |
| Rhabdomyosarcoma | 1 | 2 | 3 |
| Sarcoma | 1 |  | 1 |
| Schwannoma | 1 |  | 1 |
| Teratoma | 2 |  | 2 |
| Wilms tumor | 17 | 11 | 28 |
| Not Reported | 1 | 2 | 3 |
| **Totals:** | **76** | **97** | **173** |

The 173 patients came from eight of Cameroon’s ten regions to be treated at one of the three study hospitals. 105 (61.8%) of the patients treated came from the Northwest, with 12.4% of patients from the Centre Region and 10% from the West Region. A breakdown of patients by region can be found in **Figure 3**.

**Figure 3:**
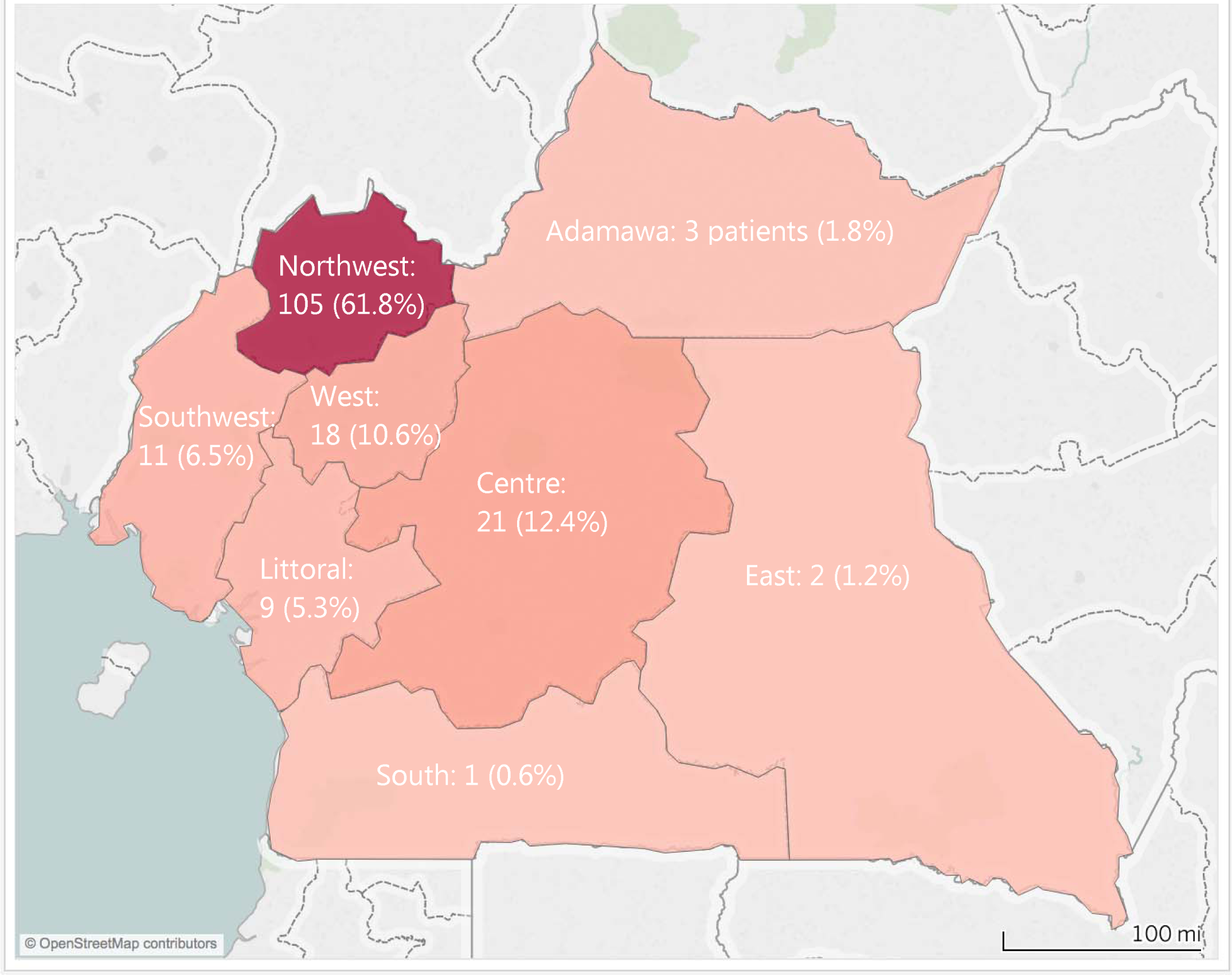
Pediatric Cancer Patients by Region.

The overall pediatric cancer incidence across all cancer types for patients’ aged 0-14 in Northwest Cameroon is 5.92 cases per 100,000 persons per year, with a 95% confidence interval of 4.79 to 7.05. The confidence interval was determined using the normal approximation to the Poisson distribution due to the large sample size and the low incidence proportion.^12^

## Conclusions

With the increasing incidence of pediatric cancers in developing countries across the world, a critical need exists to ensure that the epidemiological evidence used in practice and policymaking is of high quality. This research provides comprehensive epidemiological data on pediatric cancer in the Northwest Region of Cameroon for the very first time.

Because little existing data exists concerning the epidemiology of cancer in Northwest Cameroon, the primary point of comparison is the 2012 Yaoundé Cancer Registry study of cancer trends in the capital, which did not report pediatric cancer incidence specifically.^5^ There were 84 new pediatric cancer cases in Yaoundé in 2010-2011 as compared to the 105 new cancer patients from the Northwest Region.

There are several limitations to this study. First, a truly comprehensive dataset would be impossible to achieve for several reasons. Families often do not seek medical help until children are very ill, due in part both to the cost of healthcare and also to the difficulty of travel. Additionally, traditional healers still play an integral role in Cameroon’s healthcare infrastructure. A study conducted in part through MBH found that over 40% of children with Burkitt lymphoma were first seen by a traditional healer.^14^ Potential patients often do not even make it to a hospital that is equipped to treat cancer effectively.

A second limitation is the data imputation necessary to complete the analysis. This is connected with the occasional inconsistent reporting of demographic data, specifically with home address, height, weight, and age. Much of the data is self-reported and all of it is hand-written into a physical file, leaving considerable margin for error.

While overall cancer incidence rates are greater in high-income countries than in sub-Saharan Africa, pediatric cancer is more prevalent by proportion in Africa.^15^ Research has found that 80% of all children and adolescents diagnosed with cancer live in the developing world. These pediatric patients have significantly less access to quality care and therefore treatment than their counterparts in developed countries.^16^

While protocols and resources such as the International Society of Pediatric Oncology (SIOP) Africa and the PODC Collaborative Wilms Tumor Project exist to assist both patients and providers, considerable work still needs to be done in order to help children with cancer in developing countries achieve the same outcomes as children in developed countries.^17^ Available pediatric oncology facilities are small both in number and in size,^18^ but researchers must also build on augmenting the critical supplementary resources necessary for improving the environment of cancer care in not only the Northwest Region but also Cameroon and the entirety of sub-Saharan Africa.

## Data Availability

Anonymized data available upon request.

## Acknowledgements

This research was supported by Dr. Albert Domm, a pathologist based out of Columbia, TN. The authors thank Dr. Domm for his assistance and guidance.

The authors thank Dr. Nancy Palmer, head of the Cameroon Baptist Convention’s Institutional Review Board, for her input and assistance with the manuscript. The authors also thank Juan Antonio Castro Price, biomedical engineer, for his assistance with data collection.

## Notes

### Competing Interest Statement

The authors have declared no competing interest.

### Funding Statement

Funding was provided by the Cameroon Baptist Convention and associated physicians.

